# Access to Anti-Obesity GLP-1s for Medicare-Aged Adults

**DOI:** 10.1101/2024.03.26.24304923

**Authors:** Patricia J. Rodriguez, Vincent Zhang, Samuel Gratzl, Brianna M Goodwin Cartwright, Duy Do, Nicholas Stucky, Ezekiel J. Emanuel

**Affiliations:** Truveta, Inc, Bellevue, WA; Healthcare Transformation Institute. Department of Medical Ethics and Health Policy, Perelman School of Medicine at the University of Pennsylvania, Philadelphia

## Abstract

**Background:** Obesity is common among older adults in the US, but Medicare policies prohibit coverage of anti-obesity medications (AOMs) and may limit access to effective treatment.

**Objective:** To describe first-time prescribing and dispensing of AOM glucagon-like peptide-1 receptor agonists (GLP-1 RA) among eligible older adults, stratified by Medicare age eligibility.

**Methods:** Adults aged 60-69 with overweight or obesity and without Type II diabetes (T2D) were identified from Truveta Data. Data included EHR (encounters, prescriptions, conditions, BMI) and medication dispensing for a collective of US healthcare systems. Eligibility required an outpatient office encounter between June 2021 and January 2024 with a BMI ≥ 27, a negative history of GLP-1 RA use, and a follow-up ≥60 days later. Patients were stratified by Medicare age eligibility (60-64 vs. 65-69) and the proportions prescribed and subsequently dispensed AOM GLP-1 RA were compared.

**Results:** In total, 413,833 AOM eligible older adults were included in our cohort, with 208,067 (50.3%) Medicare-aged adults and 205,766 (49.7%) adults aged 60-64. Among eligible patients, 0.2% of Medicare-aged patients and 0.4% of patients aged 60-64 were prescribed AOM GLP-1 RA, a significant difference (p < 0.01). Among those prescribed AOM GLP-1 RA, 15.2% of Medicare-aged patients and 22.7% of patients aged 60-64 were dispensed AOM GLP-1 RA within 60 days, a significant difference (p<0.01). In both age groups, prescribing and dispensing were more common for females and those with higher BMI.

**Conclusions:** Fewer than 1% of older adults with overweight or obesity and without T2D were prescribed an AOM GLP-1 RA. New use of GLP-1 RA was significantly lower for Medicare-aged adults, compared to 60-64-year-olds, with differences occurring at both medication prescribing and dispensing stages. While coverage of AOMs is limited by many insurers, Medicare’s unique prohibition on AOM coverage may contribute to differentially lower use among Medicare-aged adults.

## Introduction

Over 40% of older adults in the US have obesity.^1^ Highly effective anti-obesity medication (AOM) glucagon-like peptide-1 receptor agonists (GLP-1 RA) are now available,^2–4^ but may be inaccessible to older adults due to Medicare policies that prohibit AOM coverage. While the Center for Medicare and Medicaid Services (CMS) recently announced some coverage of AOM GLP-1 RA when labelled for cardiovascular benefit, broader coverage of AOMs remains an open policy question.^7,8^

To explore current gaps in AOM access for Medicare-aged adults, we characterize the proportions of eligible patients aged 60-64 vs. 65-69 who were prescribed and dispensed AOM GLP-1.

## Methods

We identified adults aged 60 – 69, without Type II diabetes (T2D), who were eligible for on-label AOM GLP-1 RA between June 2021 and January 2024 using a subset of Truveta EHR and dispensing data. Eligibility required an outpatient office encounter with a body max index (BMI) ≥27 kg/m^2^ and no previous prescribing or dispensing of any GLP-1 RA (AOM or anti-diabetic medication versions). To assess 60-day initiation outcomes, we required another encounter ≥60 days later. The most recent encounter meeting these criteria per patient was used. Data were extracted on March 6, 2024.

Prescribing of AOM GLP-1 RA at the encounter was identified from EHR data, using brand names to restrict to AOM versions (liraglutide [Saxenda], semaglutide [Wegovy], and tirzepatide [Zepbound]). Initiation of AOM GLP-1 RA in the subsequent 60-days was identified from linked dispense data. We describe the number and characteristics of patients eligible, prescribed, and initiated on an AOM GLP-1 RA for ages 60-64 vs. 65-69. While age is an imperfect proxy for insurance, 94% of adults ≥65 have Medicare insurance.^9^

## Results

Our population included 413,833 AOM eligible adults without T2D, including 208,067 (50.3%) Medicare-aged adults and 205,766 (49.7%) adults aged 60-64. Among eligible patients, 54% were female, 81% were white, and 34.6% had BMI >35 (Class 2 or 3 obesity) (Table 1).

**Table 1:** Characteristics of Older Adults Eligible for AOM GLP-1 RA, by Age Group. Continuous variables reported as Median (IQR). Categorical variables reported as number (%). Black includes Black and African American. Other race includes American Indian or Alaska Native, Native Hawaiian or Other Pacific Islander, Declined to answer, Other Race, and Unknown. Overweight includes BMI 27 to <30. Class 1 obesity includes BMI 30 to <35. Class 2 obesity includes BMI 35 to <40. Class 3 obesity includes BMI≥40. *Counts for sociodemographic groups with ≤10 patients are not presented.

| Characteristic | Eligible |  | Prescribed |  | Initiated |  |
| --- | --- | --- | --- | --- | --- | --- |
|  | Age 60-64<br>N = 205,766 | Age 65-69<br>N = 208,067 | Age 60-64<br>N = 761 | Age 65-69<br>N = 495 | Age 60-64<br>N = 173 | Age 65-69<br>N = 75 |
| **Age** | 62.5 (61.2, 63.7) | 67.4 (66.2, 68.7) | 62.3 (61.1, 63.6) | 67.1 (66.1, 68.6) | 62.1 (61.0, 63.4) | 66.9 (65.9, 68.3) |
| **Female** | 110,872 (53.9%) | 111,298 (53.5%) | 607 (79.8%) | 368 (74.3%) | 143 (82.7%) | 57 (76.0%) |
| **Race** |  |  |  |  |  |  |
| **Asian** | 3,877 (1.9%) | 3,487 (1.7%) | * | * | * | * |
| **Black** | 14,959 (7.3%) | 13,118 (6.3%) | 63 (8.3%) | 35 (7.1%) | 15 (8.7%) | * |
| **White** | 162,832 (79.1%) | 170,142 (81.8%) | 605 (79.5%) | 405 (81.8%) | 137 (79.2%) | 65 (86.7%) |
| **Other** | 24,098 (11.7%) | 21,320 (10.2%) | 86 (11.3%) | 47 (9.5%) | 19 (11.0%) | * |
| **BMI** | 32.6 (29.3, 37.4) | 32.2 (29.1, 36.8) | 36.8 (33.0, 41.8) | 36.7 (32.7, 42.4) | 36.9 (33.1, 40.4) | 36.3 (33.0, 42.6) |
| **Class** |  |  |  |  |  |  |
| **Overweight** | 62,837 (30.5%) | 68,239 (32.8%) | 44 (5.8%) | 35 (7.1%) | 9 (5.2%) | 6 (8.0%) |
| **Class 1 Obesity** | 68,757 (33.4%) | 70,261 (33.8%) | 254 (33.4%) | 164 (33.1%) | 60 (34.7%) | 30 (40.0%) |
| **Class 2 Obesity** | 39,176 (19.0%) | 38,115 (18.3%) | 212 (27.9%) | 125 (25.3%) | 57 (32.9%) | 11 (14.7%) |
| **Class 3 Obesity** | 34,996 (17.0%) | 31,452 (15.1%) | 251 (33.0%) | 171 (34.5%) | 47 (27.2%) | 28 (37.3%) |
| **Comorbidities** |  |  |  |  |  |  |
| **Asthma** | 26,828 (13.0%) | 26,203 (12.6%) | 131 (17.2%) | 100 (20.2%) | 30 (17.3%) | 14 (18.7%) |
| **COPD** | 16,028 (7.8%) | 19,074 (9.2%) | 46 (6.0%) | 36 (7.3%) | 10 (5.8%) | 5 (6.7%) |
| **CKD** | 9,303 (4.5%) | 14,626 (7.0%) | 39 (5.1%) | 31 (6.3%) | 12 (6.9%) | 2 (2.7%) |
| **Hypertension** | 110,155 (53.5%) | 125,036 (60.1%) | 497 (65.3%) | 348 (70.3%) | 125 (72.3%) | 55 (73.3%) |
| **Hyperlipidemia** | 114,384 (55.6%) | 132,710 (63.8%) | 486 (63.9%) | 376 (76.0%) | 111 (64.2%) | 56 (74.7%) |
| **Major Depression** | 23,829 (11.6%) | 25,832 (12.4%) | 161 (21.2%) | 131 (26.5%) | 33 (19.1%) | 25 (33.3%) |

Prescribing of AOM GLP-1 RA was rare overall (n = 1,256 [0.3%]), but relatively more common among females (78% of prescribed) and patients with higher BMI (60.4% had BMI >35) (Table 1). Prescribing was significantly lower among Medicare-aged older adults (0.24% % of eligible patients), compared to those aged 60-64 (0.37% of eligible patients), an absolute difference of 0.13% (95% CI: 0.10%, 0.17%; p<0.01). Lower prescribing for Medicare-aged adults occurred across all BMI classes (Figure 1).

**Figure 1:**
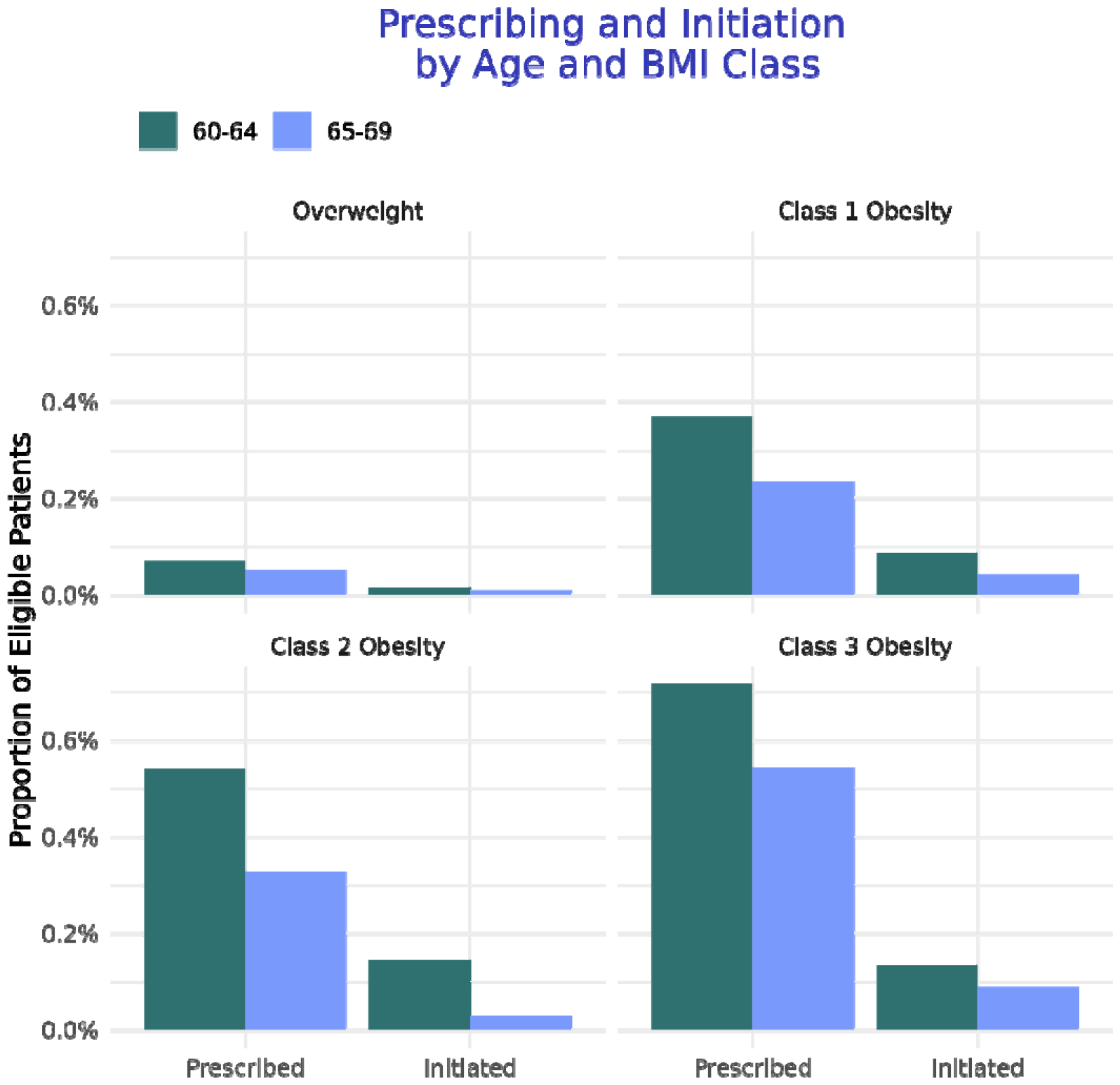
Proportion of eligible patients prescribed and initiated on AOM GLP-1, by age group and BMI class. Overweight includes BMI 27 to <30. Class 1 obesity includes BMI 30 to <35. Class 2 obesity includes BMI 35 to <40. Class 3 obesity includes BMI ≥40.

Among those prescribed an AOM GLP-1 RA, a minority filled the prescription within 60 days (248 [19.7%]). Medicare-aged adults had significantly lower rates of 60-day dispensing (15.2% of those prescribed), compared to those aged 60-64 (22.7% of those prescribed), an absolute difference of 7.6% (95% CI: 3.1%, 12.0%; p <0.01). Lower initiation rates for Medicare-aged adults were observed across BMI classes (Figure 1).

## Discussion

Prescribing and initiation of AOM GLP-1 RA in eligible older adults without T2D were uncommon overall, but significantly lower in Medicare-aged adults. Rates of new use among Medicare-aged adults were less than half those in adults aged 60-64, with differences observed across classes of BMI.

While AOM coverage is limited by many insurers, these findings suggest that Medicare-aged adults face unique gaps in access, occurring at both the medication prescribing and filling stages. Given the large number of patients eligible for AOM GLP-1 RA and high cost of these medications, increases in use to levels observed in 60–64-year-olds could have substantial budget impacts. Further research is needed to assess the health consequences of Medicare AOM coverage and expected lifetime costs – including reductions in major adverse cardiovascular events (MACE).

## Conclusion

Prescribing and initiation of AOM GLP-1 RA are lower for Medicare-aged adults, which may be due to a lack of coverage. Medicare coverage of AOM GLP-1 RA has the potential to increase prescribing and initiation substantially. Further research is needed to understand tradeoffs between increased prescribing costs and potential reductions in MACE, as well as gains in health-related quality of life.

## Data Availability

The data used in this study are available to all Truveta subscribers and may be accessed at studio.truveta.com.

